# Does visual error augmentation offer advantages during bimanual therapy in individuals post stroke? A randomized controlled trial

**DOI:** 10.1101/2025.06.04.25328824

**Authors:** Courtney Celian, Teresa Puzzi, Martina Verardi, Erica Olavarria, Federica Porta, Alessandra Laura Giulia Pedrocchi, James L. Patton

## Abstract

**OBJECTIVE:** Reaching training with error augmentation (EA) has recently shown great promise for enhancing bimanual therapeutic training, using both robotic forces feedback (haptics) and a visually distorted display elements (graphics) to amplify motor learning.

**METHODS:** Here in a two-arm, randomized controlled trial we explored the effect of visual EA alone by visually shifting the paretic limb’s cursor in the direction of error. We invited 38 chronic (> 8 months post injury) stroke survivors to practice bimanual reaching for approximately 40 minutes, 3 days per week, for three weeks.

**RESULTS:** Arm motor section of the Fugl-Meyer (AMFM; maximum score 66 points) increased an average of 2.2 and retained to a follow-up evaluation 7-9 weeks (about 2 months) later (average 1.5). Clinically meaningful increase for AMFM for chronic stroke survivors is 5.2 points. No superiority was detected due to the EA treatment, but other measures on the composite abilities (range of motion, bimanual symmetry, and movement time) showed improvements favoring EA.

**CONCLUSIONS:** While removing robot forces led to smaller gains than previous work, such touch-free bimanual therapy may still prove to be an effective inexpensive automated rehabilitation tool for wider accessibility in therapy interventions.

This study was registered at ClinicalTrials.gov (ID#NCT03300141).

## INTRODUCTION

Stroke is a leading cause of disability.^1^ Despite the many advances in rehabilitation, stroke survivors continue to experience life-long reduction in function, necessitating the growing need for effective treatment. This is particularly true for the multifunctionality of the upper extremity. Repetitive reaching exercise have increases in various measures,^2^ but available therapy time is brief, and recovery is rarely complete.^3^ Technology adjuncts in rehabilitation can aid and often enhance with less economic demand. Strategies employing knowledge of the mechanisms of neuroplasticity help reach common goals of improved range of motion, coordination, accuracy, and efficiency. Here we explore bilateral exercises enhanced by specialized form of augmented reality, error augmentation.

Our research is based upon prior studies centered on error augmentation (EA) - a technique that amplifies errors during repetitive practice sessions. This technique capitalizes on the essential role of error in neural adaptation, enabling individuals to leverage their own mistakes to trigger change.^4,5^ However, individuals with compromised sensorimotor function may have difficulty noticing error during tasks or activities.^6^ Our group and others have clinically evaluated the outcomes of EA treatment, where errors were artificially enhanced during repetitive training. This tends to effect modest improvements in learning for both healthy and individuals with a history of stroke.^7–11^ We believe this is because magnified errors lead to heightened corrections, which is a form of error-driven or *supervised* neuroplasticity.^12^ Such error-driven learning is one aspect of the neuroplasticity that is observed during skill acquisition.^8,13–17^ Quite simply, if error drives learning, then artificially enhanced error can enhance learning.

This concept of EA was tested by our group in a blinded, randomized, and controlled study where we employed the principle of EA in a collaborative therapist-participant-machine trio.^9^ As a therapist provided a movement cue to participants, an interactive robot system visually and haptically magnified their errors in real-time, without the participant’s knowledge. The EA group demonstrated significant advantages over the sham group that had similar practices but no EA. This demonstrated a clear superiority to robot-facilitated therapy in a blinded, randomized, controlled trial. This was an important result, because robotic treatments have not always shown superiority over regular practice or the typical standard of care.^18–20^

The many variations possible in this approach prompt us to explore which parts are most needed for efficacy. A follow-up study focused on bimanual tasks where the participants provided cues to themselves to determine if similar results might be possible.^10^ Tracking cues come from the participant’s own contralateral, less-affected arm, allowing for self-rehabilitation with EA.^10^ Studies on bimanual training for stroke recovery have suggested that bimanual training engages additional cortical areas and can influence the formation of new task-relevant networks and connections in a reorganization associated with restored motor function.^21–25^ An advantage of bimanual therapy is the possibility for additional “solo” training that can occur after daily one-on-one therapy has completed. This self-telerehabilitation variation again tested EA, with the additional ability to compare our prior studies with similar protocols. This second study also demonstrated a clear superiority to robot-facilitated therapy in a blinded, randomized, controlled trial.

A lingering question from previous studies is whether the robot was *necessary*, or whether a simpler setup of a tracking system might allow the illusion of distortion simply by displacing the visual feedback to look like the hand was being pushed further in the error direction. Here we present results from a visual distortion environment that shifts the subject’s cursor in the direction of instantaneous error as if it is being pushed by a robot. We included the additional challenge using both limbs to transport and balance a ball on a tray -- *TrayBall*. As before, the question was whether the observed adaptations generalize to movement ability outside the device, as similar research suggests that only 40% of adaptation transfers to motor ability after exiting the machine.^26^

Moreover, beyond the explicit rationale for the removal of robotic haptics, it is imperative to recognize the potential implications of this decision on the overall therapeutic approach. By shifting the focus from robotic interventions to a visual distortion environment, we aim to explore the fundamental principles underlying error augmentation in stroke rehabilitation while minimizing the reliance on complex technological infrastructure. This methodological shift not only allows for a more cost-effective and scalable intervention but also fosters a deeper understanding of the mechanisms driving neuroplasticity and motor learning post-stroke. Additionally, by embracing simplicity in our approach, we aim to empower both clinicians and patients with tools that are readily accessible and adaptable to diverse clinical settings and individual needs. Through this deliberate departure from conventional robotic-assisted therapies, we endeavor to pave the way for innovative, evidence-based rehabilitation strategies that prioritize efficacy, accessibility, and patient-centered care.

To test this, we again measured the effects of our training on clinical scores and hypothesized that bimanual training with EA should achieve the greatest participant gains in the arm motor section of the Fugl-Meyer (AMFM). This work was presented previously in preliminary form presenting feasibility and safety.^26,27^

## METHODS

### Setting and subjects

This two-arm, randomized controlled trial was completed at the Robotics Lab within the Shirley Ryan AbilityLab (SRALab) rehabilitation hospital. The SRALab is an inpatient and outpatient facility where patients, clinicians, engineers, and scientists meet and do research within the clinical treatment areas.

We recruited participants through our Clinical Neuroscience Research Registry and through marketing via websites and available poster/flyers. The Northwestern University Institutional Review Board (Chicago, IL) approved this study March 2nd, 2017 (STU00204661). All participants provided written informed consent, according to the Declaration of Helsinki of 1975 as revised in 2013, prior to commencing the study. The reporting of this study conforms to the CONSORT statements.^28,29^ This project was registered on clinicaltrials.gov (ID#NCT03300141).

This study was a standard two-arm blinded randomized control trial testing a treatment group receiving visual error augmentation (EA) feedback, and a control group that received an equivalent amount of practice in the same device with veridical feedback (no EA). Based on results of the previous study^,5,9,10^ we estimated that 14 subjects per group were needed to detect significance.

The target population for the study was chronic stroke participants (at least 8 months post-injury), aged 18 years or older, with a history of a single stroke event. Individuals included in the study had mild to moderate upper extremity impairment (as confirmed by an AMFM score of 15-56 out of 66 completed by an occupational therapist), and residual active elbow flexion and extension when their arm was supported against gravity. Individuals were excluded from the study if they had multiple strokes, exhibited bilateral upper extremity paresis, and/or severe sensory deficits in the affected limb. Participants were also excluded if they had severe spasticity in the arm or hand preventing movement measured as a Modified Ashworth score of 4. Individuals with aphasia that would influence their ability to perform the experiment were also excluded; this was determined by a score on the NIH stroke scale item #9 > 1. Individuals with cognitive impairment, determined by the Mini Mental State Examination <23/30, were also excluded from the study. Participants were also excluded if they were currently receiving any other skilled arm rehabilitation, participated in similar robotics interventions previously, or had other neurological/medical issues that affected their safe participation. All participants were assessed by an expert occupational therapist during a preliminary screening session to ensure participants met inclusion criteria.

Each qualified participant was randomly assigned to one of the two groups: (1) the treatment group (or EA group) (N=19), trained with error-augmented visual feedback, or the (2) control group (N=19) trained with no visual distortions. We used a block randomization method to ensure an equal number of participants presenting with mild and moderate stroke symptoms were assigned into both groups. Participants were randomized 4 at a time based on their AMFM scores assessed during the initial screening visit. The blocks had an inclusion range of six AMFM points. Every other participant in the same group of 4 was assigned to a group such that they balanced that group, otherwise the assignment was random.

We considered an improvement in the arm motor section of the Fugl-Meyer (AMFM) score as the primary outcome measure for this study. An improvement equal or greater than 5.2 is commonly believed to be clinically meaningful.^30^

### Apparatus

To implement the study design, we developed an immersive three-dimensional graphics display system called the LookingGlass. The LookingGlass is a stereographic 3D visual display with a semi-silvered mirror that allows viewing of a large visual field and free movements of the hand in front of the user (Figure 1A). The LookingGlass was made in-house at the SRALab Robotics Lab. Stereographic liquid crystal shutter glasses synchronized separate left and right eye images with the TV resulting in the perception of depth. Mobile arm supports attached to an armless, stationary chair were used to assist the participant in maintaining a partially flexed shoulder position and to avoid fatigue during the reaching exercise block. Used in conjunction with the LookingGlass graphic display was a Leap® *(UltraLeap, Mountain View, CA, USA)* tracking tool, which allowed for a full field of view to track limb movement (Figure 1C). An in-house made orthosis was used to assist the Leap® tracking tool if participants had difficulty with obtaining full digit extension and abduction during the reaching movement (Figure 1B).

**Figure 1.**
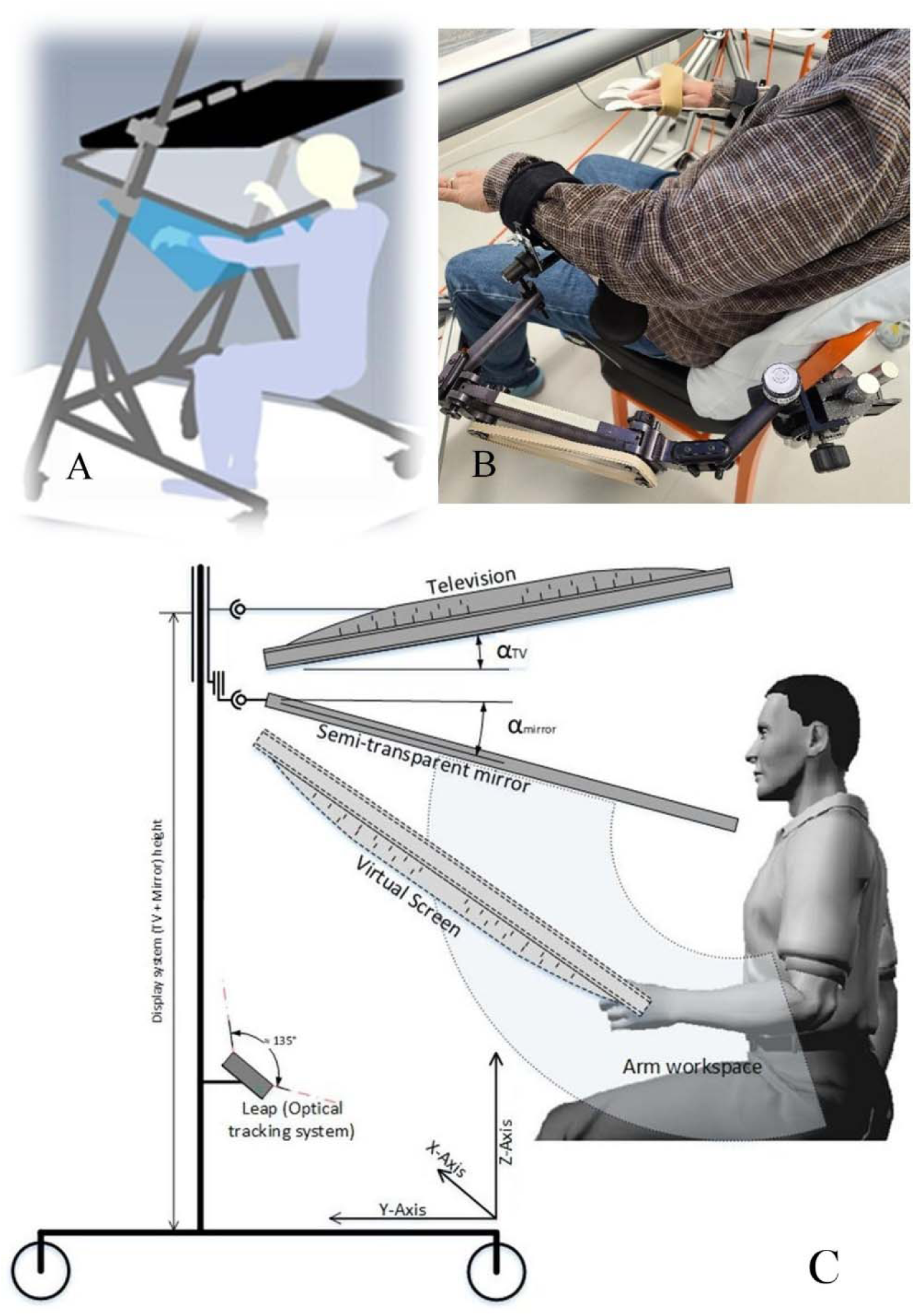
(A) Schematic of LookingGlass augmented reality system. (B) Apparatus in the clinical setting with a subject reaching in the augmented reality environment assisted by mobile arm supports and an orthosis (right hand). (C) The Leap® Motion Controller hand-tracking sensor is positioned parallel to the ground on a rigid support under the hands of the subject.

### Protocol

We asked chronic stroke survivors to practice a bimanual transport task in an augmented reality environment for approximately 40 minutes (excluding setup and breaks for rest), three times a week, for three weeks. During the treatment sessions, participants were seated in a stationary chair with both arms (hemiparetic and unaffected) supported by a gravity balanced orthosis, with their arms occluded by the screen (Figure 1B). Participant’s hand position was tracked by the Leap® tracking tool and displayed on the augmented display as two blue toy jacks (Figure 2A). In the augmented display, the participants controlled the two toy jacks with their left and right hands respectively. Participants were instructed to move the toy jacks towards the green sphere targets (Figure 2B), while simultaneously maintaining level positioning between their two hands, to keep a ball from rolling off a tray (Figure 2C).

**Figure 2.**
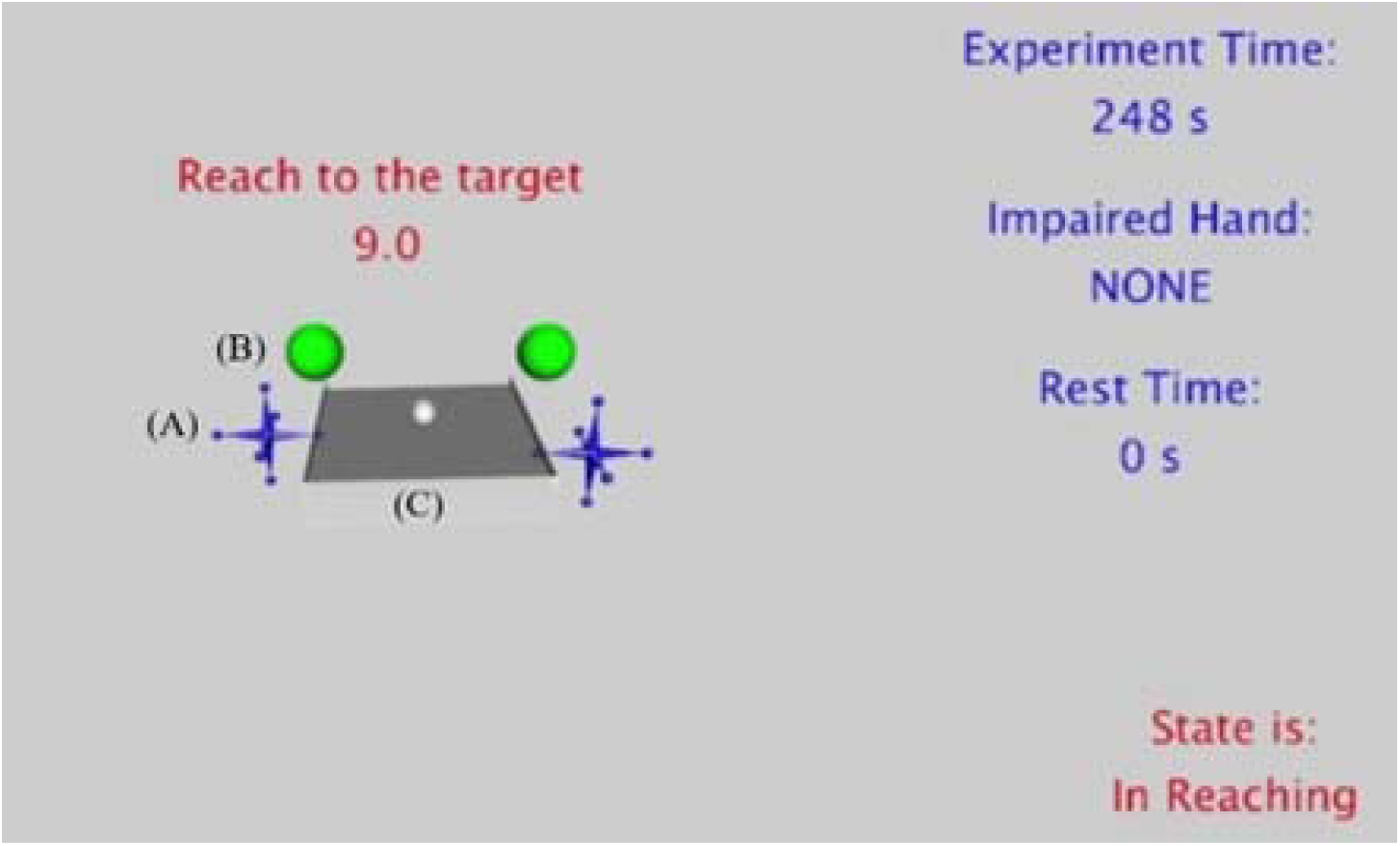
TrayBall software view. (A) The blue jacks represent the position of the hands tracked by the LEAP®. (B)The green balls are the targets that need to be reached by the participants, (C) while maintaining symmetry of hand position to balance a white ball on a gray tray.

Each treatment visit was approximately 75 minutes. It began with a 20-minute setup to position the participant and set up the apparatus, then seven, 6-minute blocks of targeted reaching trials followed by 2-minutes of rest (Figure 3B). Extra rest was taken as needed such that the subject reported little to no fatigue. Targeted reaching trials involved attempts to reach from a rest position above the centers of the thighs out with both arms to one of 15 target pairs, and then stopping for at least a half-second. Participants had 10 seconds to make this motion, at which point the system cued a return to the starting point and proceeded to the next motion. During the return to the starting point, the ball was frozen and therefore did not require tray balancing. The targets were spaced evenly on an invisible quarter sphere in front of the subject.

**Figure 3.**
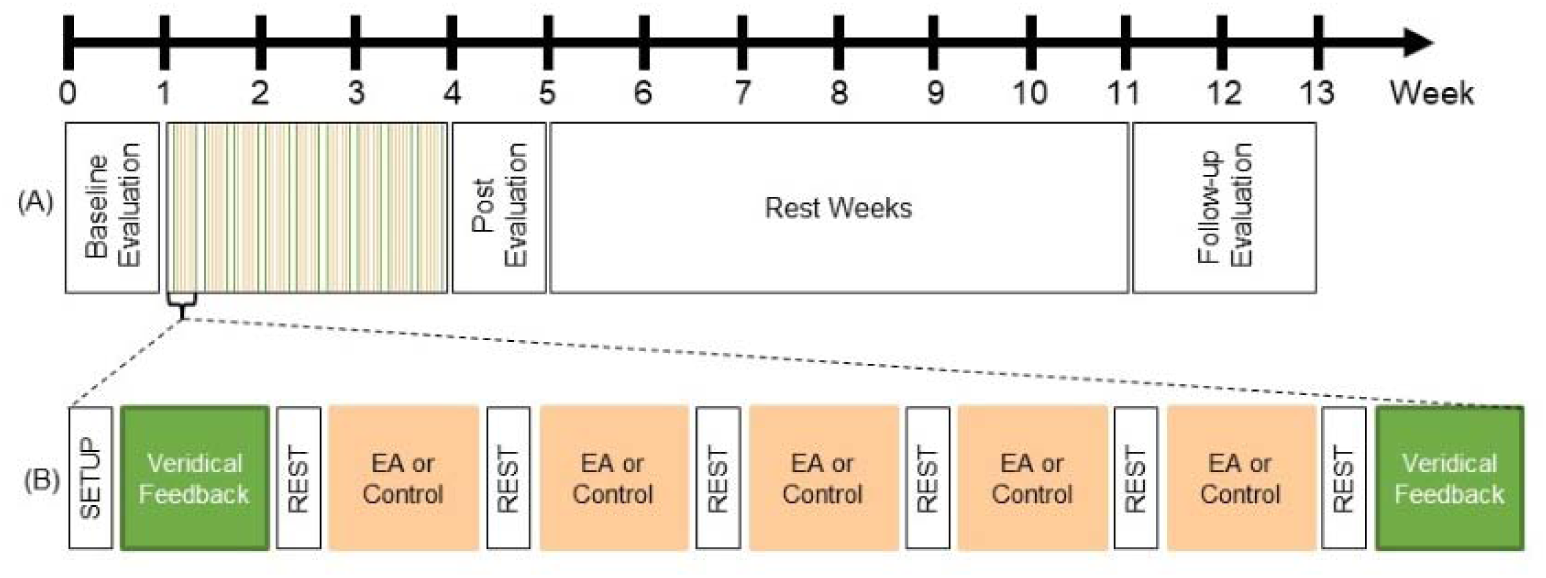
(A) Training protocol timeline. Treatment visits to the lab lasted 3 weeks with three visits per week, and clinical evaluations occurred in week 0, 5 and 12. (B) Outline of each treatment session. Each visit consisted of seven 6-minute blocks (orange) where participants completed the reaching treatment with either EA feedback applied (treatment group) or veridical feedback (control group) with 2-minute rest breaks (white), with an initial and final block where movement ability was assessed (green) with veridical feedback applied.

If subjects were at least 70% successful in their attempts to reach the targets on any block, the targets were moved 0.05 m away from the rest position for the following blocks. If less than 30% of the attempts were successful, the targets were moved back 0.05 m closer to the rest position. This was meant to probe participants’ range of motion as well as consistently provide them with an adequate level of challenge, in accordance with the challenging point theory.

Both the control and the treatment group (i.e., the group that received visual EA training) had the same amount of practice in three weeks of training with three sessions per week (nine sessions total). Each session was 50-60 minutes including rest time. The EA treatment involved visual distortions applied at each instant of time by taking the error vector, defined as the difference in position between the participant’s wrists, and visually magnifying the vertical component by a factor g of 1.3 on the paretic side.

As our primary outcome, participants were evaluated by a blinded therapist evaluator with clinical measures up to one week prior to treatment (pre), four to seven days after the end of treatment (post), and in a follow-up performed seven to nine weeks post-treatment (Figure 3A). The only exception was one participant whose return for their follow up occurred three years later due to researcher oversight.

### Outcome Metrics

Our primary outcome measure was the arm motor section of the Fugl-Meyer (AMFM) to evaluate performance-based impairments, with the maximum score being 66 points.^30,32,33^ We recorded the AMFM score at three stages during the experiment. First, we obtained scores during screening and pre-evaluation and then averaged them to establish a (1) baseline value. Then, we recorded the score during (2) post-evaluation, after the participant completed all 9 treatments. Finally, we obtained the (3) follow-up scores, 7-9 weeks (about 2 months) after post evaluation. We then compared the post-evaluation and follow-up values to the baseline ones to reveal whether training with the interface resulted in significant clinical improvement in AMFM scores. We then assessed participants’ task performance by a *Composite Error,* a comprehensive analysis of kinematic performance using our technology. Composite Error was sum of three normalized error metrics: First, range of motion deficit,

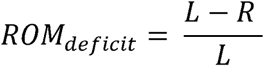

where *L* was the length of the participant’s arm and *R* is the maximum distance between the rest position and the most peripheral target. Second, vertical hand asymmetry,

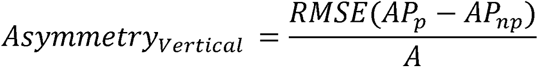

where *AP_p_* is the arm position of the paretic limb, *AP_np_* is the arm position of the non-paretic limb, and *A* is the maximum asymmetry between the left and right limb. Third, average movement time,

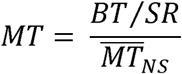

where *BT* was block execution time, *SR* was number of successful reaches, and 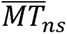 was the average movement time for non-stroke patients. All three metrics were normalized with a minimum of 0 and a maximum of 33.33 representing the largest error observed, then summed to form a *Composite Error* metric with a maximum of 100.

Other tertiary outcome measures collected at pre, post, and follow-up included: (1) the Action Research Arm Test (ARAT), a 19-item evaluation of upper extremity functional performance, where a higher score indicates more movement function (a maximum score of 57).^34^ (2) Wolf Motor Function Test (WMFT), a 17-item quantitative measure of upper extremity ability through unilateral and bimanual functional activities where a higher score indicates more movement proficiency (maximum score of 75).^35^ (3) Grip strength was assessed using a hand-held dynamometer (cut off for disability < 62 lbs. for Male, <41 lbs. for Female),^36^ and (4) The Box and Blocks Test (BBT) evaluated unilateral gross motor skills by measuring the number of blocks moved from one side of a box to another within 60 seconds.^37^

### Statistical Analysis

To test the effect of the visual EA treatment across time, we ran a non-parametric generalized linear mixed-effects model with two factors: group (EA-treatment and control) and time (pre-, post-evaluation, follow-up). The threshold for significance was *α* = 0.05. Non-parametric approaches were employed if Chi-square goodness-of-fit revealed that distributions were non-normal. We used the SPSS software *(SPSS Inc. Chicago, IL, USA)* as well as MATLAB (*MathWorks. Natick, MA, USA*).

To determine the sample size needed to reach statistical significance, we ran a simple power analysis on the simple two-arm clinical trial data from prior study.^10^ Based on a power > 80%, two-sided type I error of 0.025 (adding to 0.05) and an estimated variance of 2.5^31^ we obtained a sample size of 11 subjects per group, with an estimate of 10% participant dropout and a safety factor of 1.25, we arrive at 15 individuals per group, totaling 30 in two groups.

This study was registered at ClinicalTrials.gov (ID#NCT03300141).

## RESULTS

In this two-arm randomized clinical trial, 69 participants were screened for inclusion/exclusion criteria (Figure 4) from March 2017 to December 2021. Of those assessed, 27 participants were excluded for not meeting the inclusion criteria, and 4 declined to participate. Thirty-eight participants were randomized into two groups: (1) 19 participants in the treatment group where visual distortion (EA) was applied; (2) 19 participants were assigned to the control group with no EA applied (veridical feedback). Both the treatment and the control group had three participants each withdraw from the study for personal or unrelated medical reasons. The treatment group had 16 participants that completed the post and follow-up assessment periods for the clinical analysis, while only nine for the kinematic analysis (seven participant’s data were lost due to technical failure). The control group had 15 participants that completed all assessments for clinical analysis, while 12 for the Composite Error analysis (one participant’s data was lost due to technical failure); one participant was unavailable and lost to follow-up. One participant from the control group was not followed up until three years later, but their data was included in all analyses.

**Figure 4.**
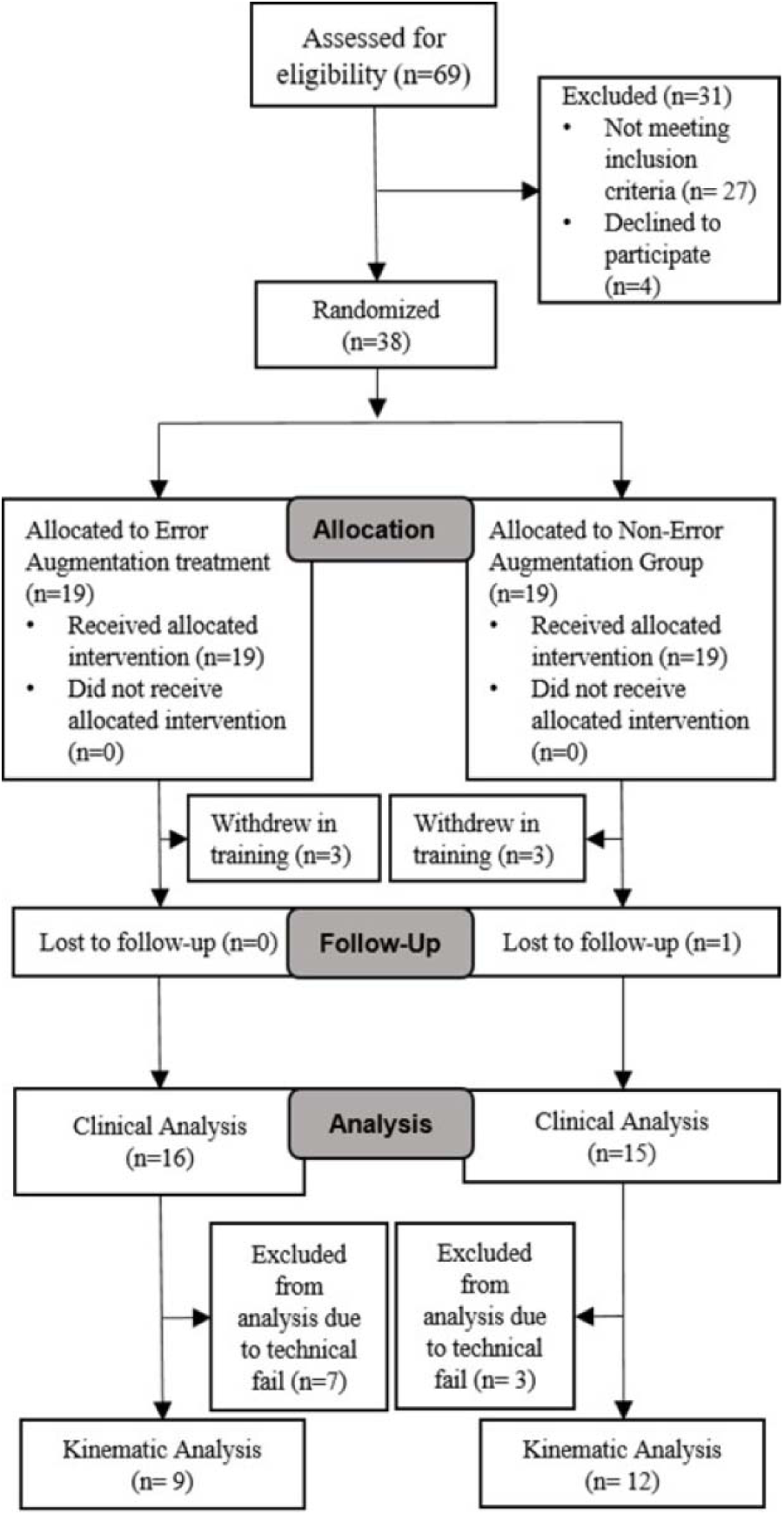
Participant recruitment flow diagram, based on the CONSORT^29^ protocols.

Demographic information of all participants in the treatment and control groups who completed clinical and kinematic performance analysis can be found in Table S1 (Supplementary Materials) of the supplementary section. Individual subject information for all participants included in clinical and kinematic data analysis can be found in the supplementary section in Table S2. Technical failures, detected only after study completion, led to the loss of kinematic performance data for 10 participants, who were excluded from the analysis (Figure 4; see Table S2, Supplementary Materials).

Normality was assessed using the Shapiro–Wilk test, which indicated that both the differences for each group were normally distributed (p = 0.6, p = 0.3 and p = 0.1, p = 0.9, respectively). We observed improvement in the clinical outcome, AMFM, across time for both groups (p=0.005, F=5.612). However, our hypothesis that EA treatment would show superior improvement was not supported (time × group interaction p=0.069, F=2.772). Interestingly, average gains seen in the treatment group were significant by post-evaluation, and the controls were not (Fig. 5). We see this as underpowered evidence suggesting that some individuals may respond better to this treatment (Fig. 5). Nevertheless, both groups improved less than 5.2, the minimal clinically important difference.^30^ We also did not detect superiority if subjects were divided into mild (AMFM > 24.5) and severe impairments, as has been done by others^19^ (mild: time × group interaction p = 0.887; severe: p=0.658).

**Figure 5.**
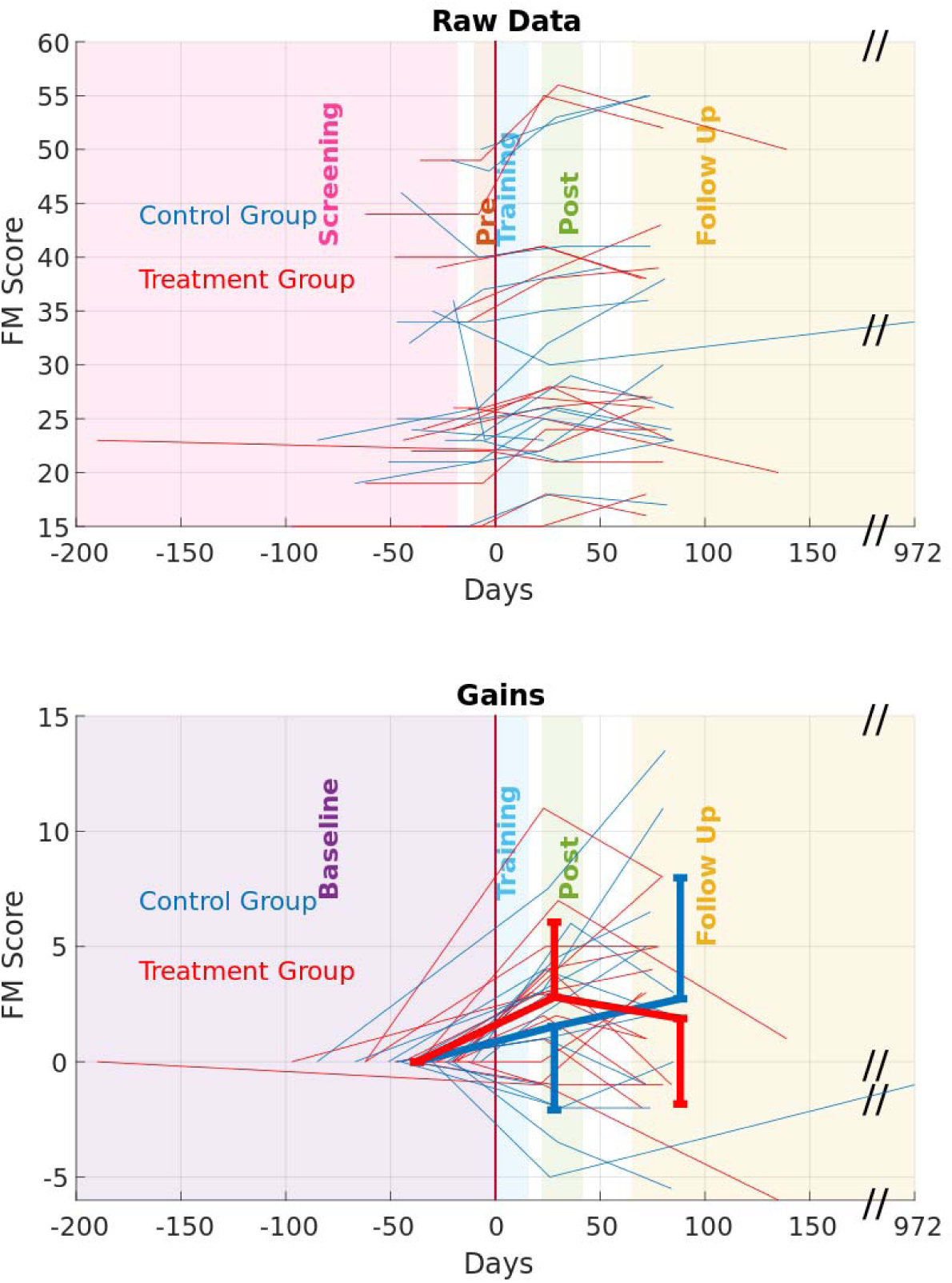
The primary outcome measure, AMFM, failed to show superiority for our customized EA treatment. Associated gains (bottom) were pooled for each group for the major regions of time and shown as thicker lines. Wings indicate variance across each treatment group. For these, time was aligned to each subject’s initial training session (time zero).

The secondary metric, *Composite Error*, demonstrated a benefit to our training by reducing between groups (p=0.031, F= 3.70). However, EA treatment did not show superior improvement (time × group interaction, p=0.575, F= 0.56), despite this group exhibiting the greatest error reduction (see Figure 6, Lower figure, red bars). We failed to notice any differences by separating out the three components of composite error. We also inspected correlation between changes of Composite Error and AMFM (r=0.2). Correlation between post-evaluation and follow-up (r=-0.35). This low correlation suggests that technology may measure different aspects of improvement in ability and function.

**Figure 6.**
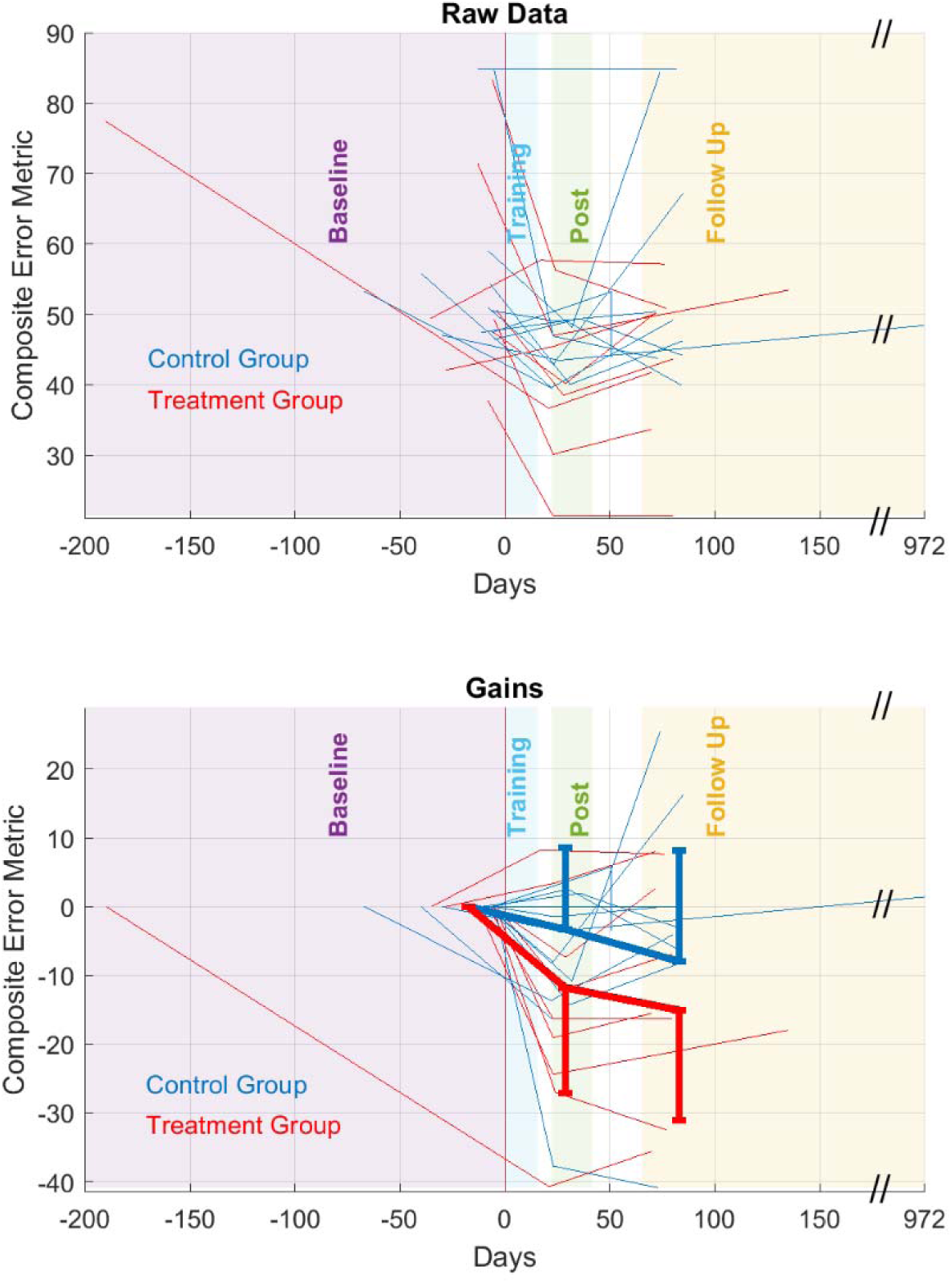
Using the same format as Figure 5, a secondary metric from our technology was our composite error, which normalized speed, accuracy, and difficulty level. This was more sensitive to changes but did not show a significant advantage for the treatment group.

We failed to notice any difference between groups from training through final follow-up visit in ARAT (p=0.48), WMFT (p=0.70), grip strength (p=0.71), and BBT (p=0.41). See figure S1 (Supplemental Materials).

## DISCUSSION

In this two-arm blinded randomized control trial, the principal objective was to explore and ascertain the efficacy of error-augmented learning (EA) integrated with visual distortions concerning motor learning and performance in a bimanual task. The primary outcome was assessing the effectiveness of these interventions, utilizing the arm motor section of the Fugl-Meyer (AMFM). The AMFM outcome did not show significant contrast between the treatment group (visual EA), and the control group. However, all subjects improved in a general metric that inspected a composite of three aspects of movement error (range of motion, vertical hand asymmetry, and average movement time) provided slightly clearer results, where the treatment group showed a decrease in Composite Error.

One possible reason for non-significance of this study, compared to our previous study, is that the special treatment (EA) was only applied to vision, rather than haptic plus visual EA.^10^ This is an important finding as it suggests that functional motor learning only occurs when the participants’ visual and haptic environments are augmented, not just visual. Consequently, the comparatively weaker outcomes observed in this study may be attributed to the fact that the treatment group received solely visual feedback without the concurrent inclusion of haptic feedback.

One subject did not complete the follow-up until three years later. While we acknowledge the deviation of methodology (follow-up period was 7-9 weeks post-treatment), we opted to include the subject in our analyses to ascertain potential insights gained from longitudinal analysis. It is also worth noting that the number of subjects in the study was relatively small, and the study was powered to detect a difference between groups in a short amount of time (three weeks of treatment and three visits per week). Hence the detected effects are likely not the full magnitude that might have been seen in a full course of treatment (up to 8 weeks). Therefore, it remains to be seen whether other implementations of bimanual EA may lead to better outcomes and insights on responsiveness. Nonetheless, training improved average AMFM scores in both groups. Detectable differences from EA treatment (vs controls) may emerge in more sustained treatment regimens or with a more focused cohort of subjects better capable of engaging in such therapy.

Error Augmentation holds promise as a rehabilitative technique to be used in conjunction with typical treatment, but the underlying neurological mechanisms that make some individuals respond to EA are still not clear. This study sheds light on the sensory sources of learning if one considers that weaker results have been gained when we remove the haptic aspects and simply distort vision. While we have found in previous studies that a combination of haptic and visual EA feedback resulted in superior outcomes to controls in arm and hand abilities,^10^ here we provided only visual feedback, which led to similar improvement, but we could not distinguish the superiority of EA. Consequent added sensory sources that are part of haptic EA (cutaneous and proprioceptive) may lead to superior recovery of motor ability.

This study differs from previous EA studies^5,9,10^ in several ways, including the replacement of free exploration with a tray balancing task, the addition of the bimanual component, and the removal of robot forces (haptics) with only hands free. Additionally, in this study the error is augmented in only the vertical direction for tray balancing, because the Z-axis is the most relevant for post-stroke patients as it represents the most challenging direction for movement. This choice was made to keep the exercise simple for patients, as overly complex tasks could reduce attention and engagement.

Error augmented learning treatment may not always be the most effective for all training conditions, and other forms of guidance may provide better results.^2–6,8,9,13^ It remains to be seen which of these factors might be the reason for the lack of clinical superiority for EA treatment seen in this study.

Our secondary measure, while not validated, is more comprehensive in assessing a broad, balanced metric that does distinguish EA as superior. While we failed to detect significant contrast in the effect of treatment (timeXgroup), we did detect the significant change only for the EA treatment, and the most dramatic changes were in the visualization of the Composite Error (Figure 6). It remains to be seen whether such metrics, when objective, appropriately normalized, and comprehensive, may offer a more sensitive measure of motor deficit than their clinical counterparts.

Our tertiary metrics, using validated clinical measures, did not detect significance between groups when looking across training through final follow-up visit. Interestingly, the treatment group demonstrates a wider spread of change in scores from pre-evaluation to follow-up for ARAT, grip strength, and BBT (fig. S.1). This variability within the treatment group suggests potential individualized responses to the intervention, warranting further investigation into personalized treatment approaches. This work represents only our analysis of *a priori* questions in the context of this completed study. Numerous other clinical metrics and motion-tracking data within this extensive data set await exploration and investigation, such as smoothness or lateral error. We anticipate that shared data from this study can be made available to the community for further mining and novel questions in the years to come.

With this study, we aimed to explore if error-driven learning with visual distortions resulted in improved motor learning and performance. While previous work used robotics, we restricted the EA treatment to be only visual, using an augmented reality display. While this visual EA training does not have comparable results to haptically implemented EA on upper extremity rehabilitation, it does offer a simple, inexpensive, and robot-free alternative in the clinic for the chronic stroke subject that leads to improvement in function. Expanding the scope of analysis through longer-term interventions involving a larger participant pool holds the potential to provide a more nuanced understanding and differentiation of this specialized form of EA treatment.

## AUTHOR CONTRIBUTIONS

CC, MV, FP, ALGP, and JLP have collectively played pivotal roles in shaping the study’s conception and design. CC, TP, MV, FP, and JLP have actively participated in data interpretation. The acquisition of data and its subsequent analysis involved the collaborative efforts of CC, MV, FP, and JLP. The manuscript drafting and critical revisions were conducted with significant contributions from CC, TP, MV, EO, FP, ALGP, and JLP, ensuring the inclusion of substantial intellectual content. All authors have provided their unanimous final approval for the version intended for publication.

## ACKNOWLEDGEMENTS

We thank Eyad Hajissa, Felix Huang, Robert Kenyon, Francois Kade, Kelly Thielbar, Yazan Abdel Majeed and Piper Hansen for their earlier help on this project. We also thank the Robotics Lab group at the Shirley Ryan AbilityLab for their insights and comments as this project has developed.

## DATA AVAILABILITY STATEMENT

The data sets generated during and/or analyzed during this study are available in our *Trayball Data repository*, www.dropbox.com/sh/atcljlhvg6wmsz5/AABlWcLkccAXnbOMaBbRT73ta?dl=0

## SUPPLEMENTARY MATERIALS (APPENDIX)

**Table S1.**
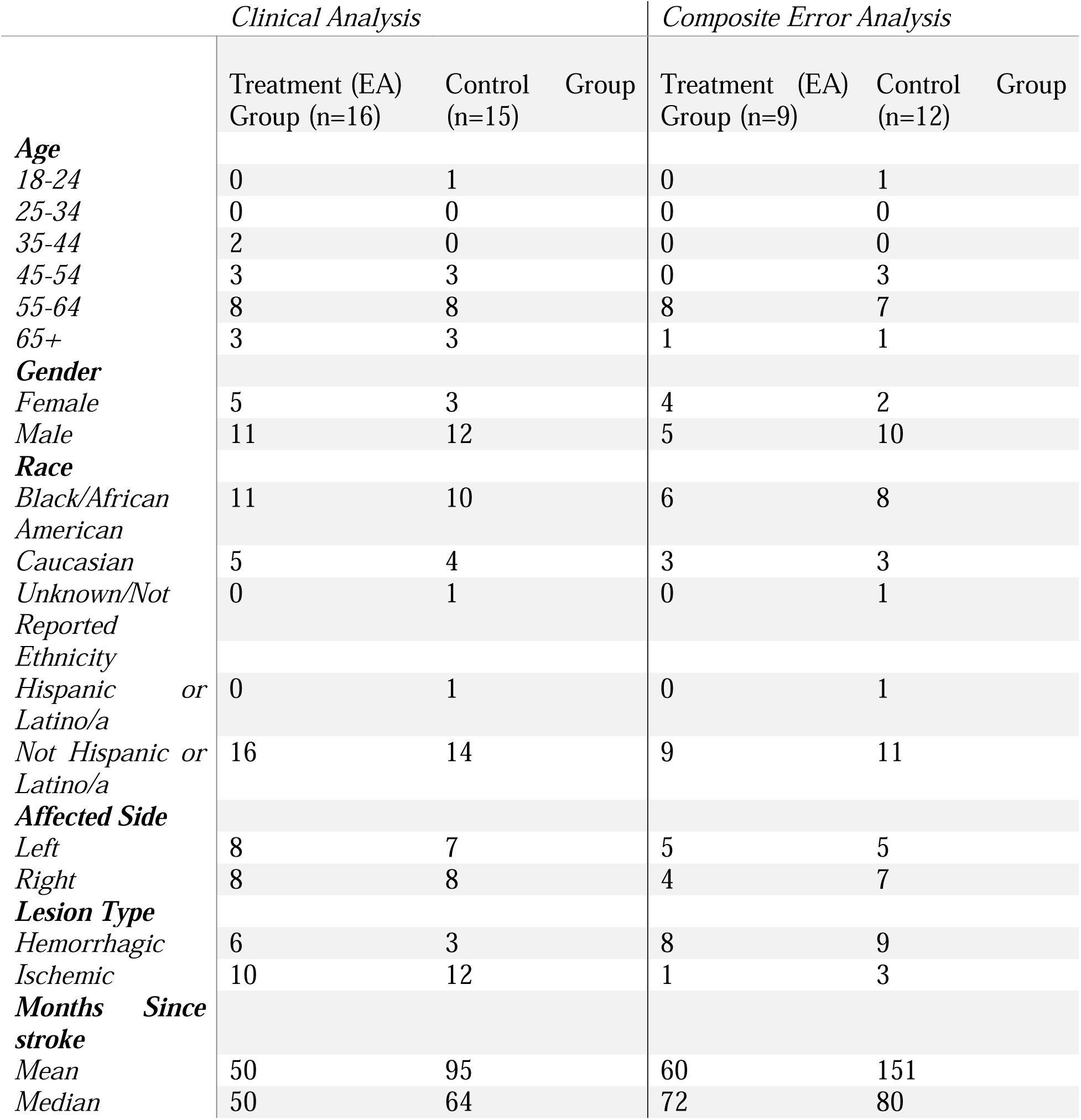
Participant demographics, and stroke information of participants included in the clinical and kinematic data analysis.

**Figure S1.**
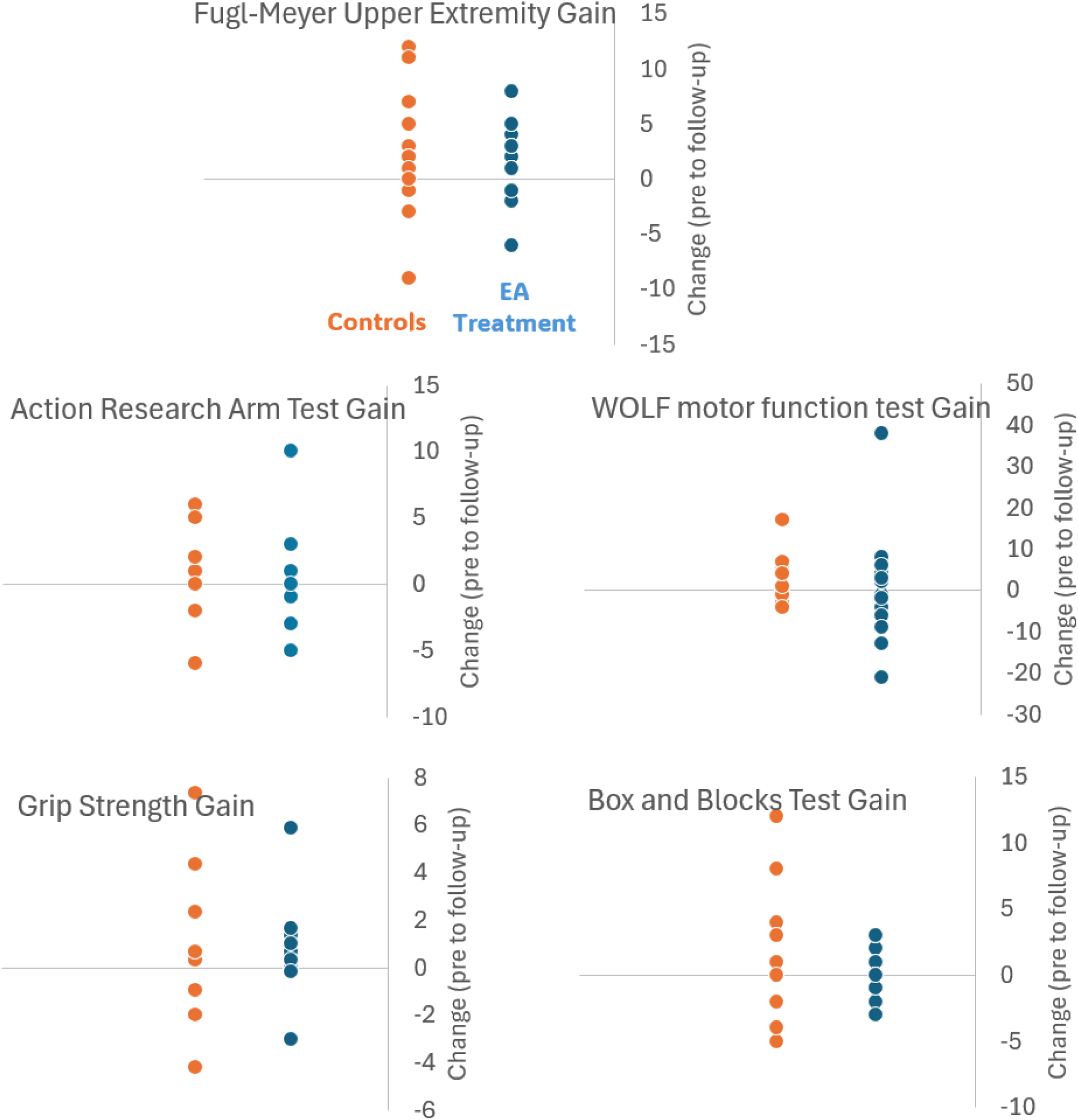
Change in clinical measures across the training through final follow-up visit.

